# CORELSA - Remote stethoscope system for fast and standardized auscultations of large numbers of patients with respiratory syndromes

**DOI:** 10.1101/2024.09.10.24313299

**Authors:** Kris Ides, Arne Aerts, Rens Baeyens, Niels Balemans, Tom Bosschaerts, Jonathan Cazaerck, David Ceulemans, Fons De Mey, Wouter Jansen, Robin Kerstens, Dennis Laurijssen, Eline Lauwers, Anthony Schenck, Girmi Schouten, Ralph Simon, Thomas Verellen, Erik Verreycken, Philippe Jorens, Stijn Verhulst, Walter Daems, Jan Steckel

## Abstract

One of the major impacts of the current COVID-19 pandemic is the immense strain that is being put on the intensive care units. As a typical SARS-CoV-2 infection often leads to severe acute respiratory syndromes, one of the most basic essential examinations is lung auscultation. There are different categories of lung sounds that can be assessed with a stethoscope like crackles, wheezes, rhonchi and coarse breath sounds. Each of these malicious lung sounds contribute to a clinical assessment and follow-up of the patient’s airways and lungs and can indicate the development of pneumonia. Therefore, auscultation on a regular basis is essential, but it also takes a lot of time for the physicians. Moreover, because of the risk of infection for health care workers, it is becoming increasingly difficult or sometimes impossible to safely use a stethoscope. Here we introduce a remote stethoscope system that can help to reduce the workload of physicians and nurses, diminishes the risk of contamination and leads to more standardized auscultation, which can also be stored and reevaluated. We provide a detailed description of our system, which consists of parts that are low cost, easily available in most parts of the world, or can be 3D printed. We provide instructions and an open source software stack to run such a system for a large number of patients which can be remotely auscultated from a central computer. We believe that the system described in this paper can allow various institutes to replicate the system, and increase the safety and reduce the overall workload in the intensive care units around the world.

## I. Introduction

THE stethoscope is one of the basic diagnostic tools in medicine and essential for the assessment and follow up of patients with respiratory syndromes[1]. While most auscultations are still conducted with traditional stethoscopes, the use of electronic or digital stethoscopes is increasing as they offer some major advantages [2]. One major advantage is that auscultation recordings can be stored, which makes it possible to reanalyze the recordings and visualize them (e.g. a spectrogram representation). Signal processing techniques, like noise cancelling [3] increases the quality of auscultation recordings.

Computer-Aided Lung Sound Analysis can further improve the diagnostic value of auscultation recordings as they are not limited by the sensitivity of the human ear [4,5]. Databases of auscultations recordings allow extraction of diagnostic sound features using machine learning frameworks such as convolutional neural networks [6]. Moreover, digital stethoscopes have already proven useful for auscultation of COVID-19 patients [7,8]. As severe courses of COVID-19 are often associated with respiratory syndromes, auscultation on a regular basis is crucial to observe the progress of the patient. However, because of the risk of infection for health care workers, it is becoming increasingly difficult to safely use a stethoscope and therefore it was already suggested to replace stethoscopes with other diagnostic devices [9]. With some of the protective gear health care workers are using, e.g. “full face” caps, it becomes virtually impossible to use a stethoscope efficiently while working safely. Another factor against conventional auscultations is that they are time consuming and hard to conduct with the increasing numbers of patients during the COVID-19 pandemic. Remote stethoscopes could be a solution for this problem and there are some remote stethoscopes available, mainly from three providers: Thinklabs, StethoMe [10], and Strados RESP technology [11]. Only the latter is designed to make remote auscultations of more than one patient at a time but this device is probably not available in large quantities and is expensive. Therefore, the widespread applicability of these systems is not readily feasible during the current COVID-19 crisis.

In this paper we describe the CORELSA system, which stands for COsys-lab REmote Lung Sound Analysis. As the name implies, CORELSA is a remote stethoscope system which can contribute to solving the aforementioned problems. Furthermore, we show how to build a network of connected remote stethoscopes using off-the-shelf components and some 3D printed parts. The network of stethoscopes can be controlled from a central computer allowing auscultations of large numbers of patients without the necessity to have direct contact with them. Along with this study we publish a GitHub repository with step by step guides, and all the necessary software components needed to run the system.

## II. Methods

### Overall workflow

With the system we propose, CORELSA, it becomes possible to conduct remote auscultations of patients without getting in direct contact to them (Fig. 1a). The system consists of one or more auscultation units that are mounted near the patients and to which digital stethoscopes (up to three for each bedside unit) are connected (Fig. 1 b,c). The auscultation units communicate via over a standard TCP/IP network (ethernet or Wifi) with a central computer from which the physician can remotely acquire and analyze auscultation recordings (Fig. 1a). Other features include scheduled measurements, and the possibility to add diagnostic remarks to the auscultation files, which allow the creation of automated machine learning models in a second stage. The labelled data from COVID-19 patients allows the implementation of machine learning models which can rely on the labelled data as input into the training phase. The CORELSA system has no hard limit to the number of bedside auscultation units that can be connected to the network.

**Fig. 1.**
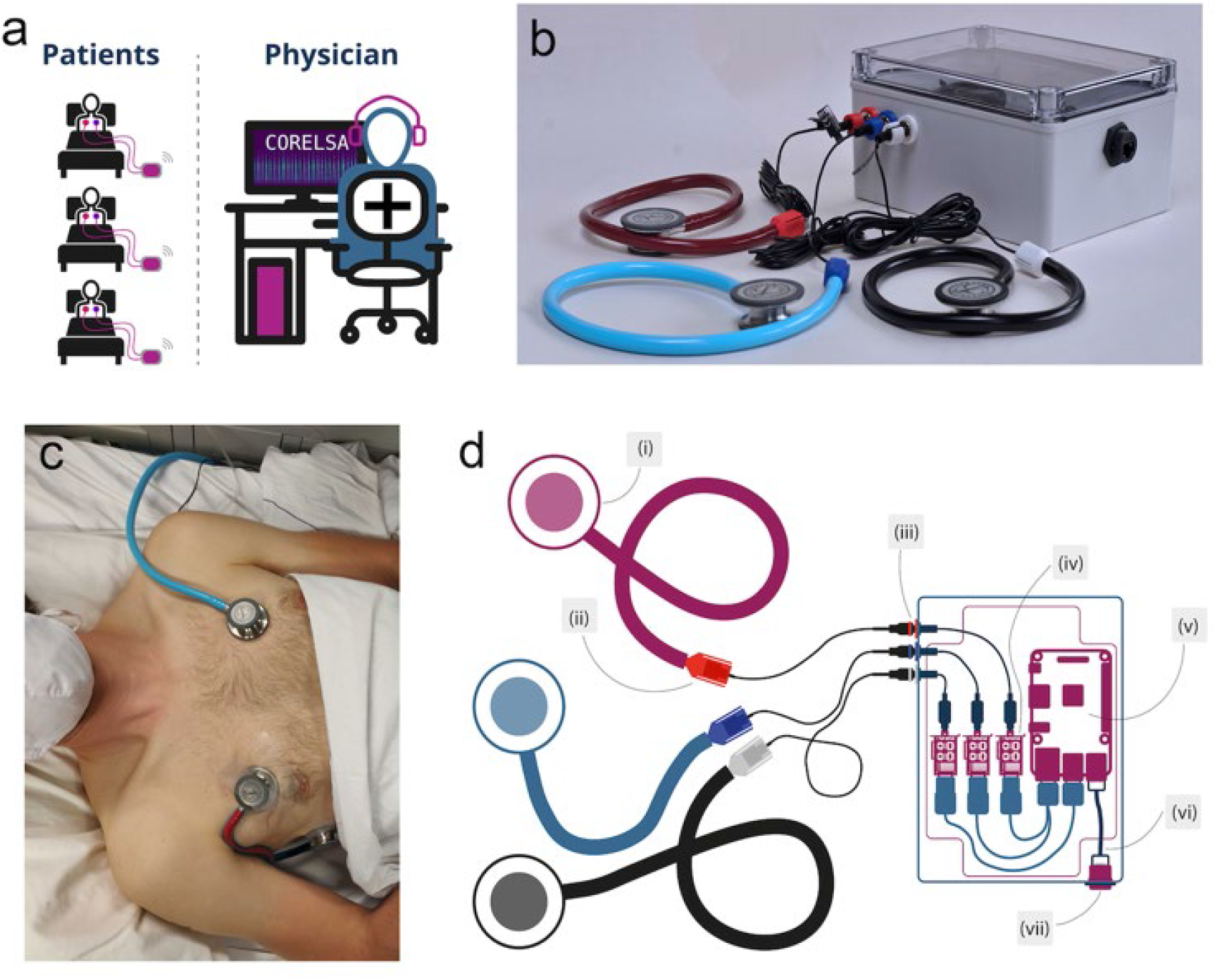
The CORELSA remote stethoscope system. **a**, Scheme of the system, auscultation units are mounted next to the patients and stethoscopes are attached to their bodies. The auscultation units can be controlled via a central computer from which the physician can remotely acquire, schedule and analyze auscultation recordings. **b**, Image of one auscultation unit with three custom made digital stethoscopes. **c**, Image of a patient’s chest with the stethoscopes fixed with medical tape on the following positions: left anterior, 3rd-4th intercostal; right anterior, 3rd-4th intercostal; right lateral, mid-axillary. **d**, Overview scheme of one auscultation unit: (i), Standard Littmann Classic III stethoscope or any other comparable stethoscope (ii), 3D printed snap-in adapter with a SM1 Sharkoon microphone. (iii) color coded microphone jacks. (iv), Renkforce 7.1 Sound card. (v) Raspberry Pi, (vi) Ethernet cable. (vii), Power over ethernet connection to provide power and connection to the network.

## III. Results

### Digital stethoscopes

We built digital stethoscopes using industry standard Littmann Classic III stethoscopes and Sharkoon microphones (Fig. 2a), which are connected via 3D printed connectors (Fig. 2b). We experimented with different adapter designs and tube lengths. Our final design was a snap-in adapter into which the microphone is inserted. The snap-in adapter itself is then inserted into the stethoscopetube; this design provided the best sound quality. A first adapter that was directly attached to the stethoscope head via several 3D printed parts (see Suppl. Fig. 1) had less gain, a noisy signal and was more complicated to produce. Another important factor in the design of the adapter is that the sound channel has to stay sealed, to avoid a loss in sound pressure level and also the recording of noise from outside. Therefore, we designed an adapter that consists only of one part, where the microphone can be snapped in and is pressed against the sound channel (Fig. 2b). This adapter is then connected to the plastic tube of the stethoscope (Fig. 2c). To determine how the tube length influences signal transmission, we measured the magnitude of the transfer function of our custom-made digital stethoscopes with different tube lengths. We excited the stethoscope membrane with linear upward frequency chirps and used a Power Spectral Density (PSD) estimator to determine the magnitude of the frequency response of the digital stethoscopes (Fig. 2d). For longer tube lengths standing wave patterns generate resonance nodes, while shorter tube lengths show fewer resonance nodes with only two nodes and a quite flat frequency response for a tube length of 15 cm. However, it should be noted that the resonance modes are relatively modest, leading us to believe that the effective tube length is not extremely important. Furthermore, due to the modest resonances in the transfer function, it is relatively easy to compensate for these non-flat transfer functions through a linear filter in post-processing.

**Fig. 2.**
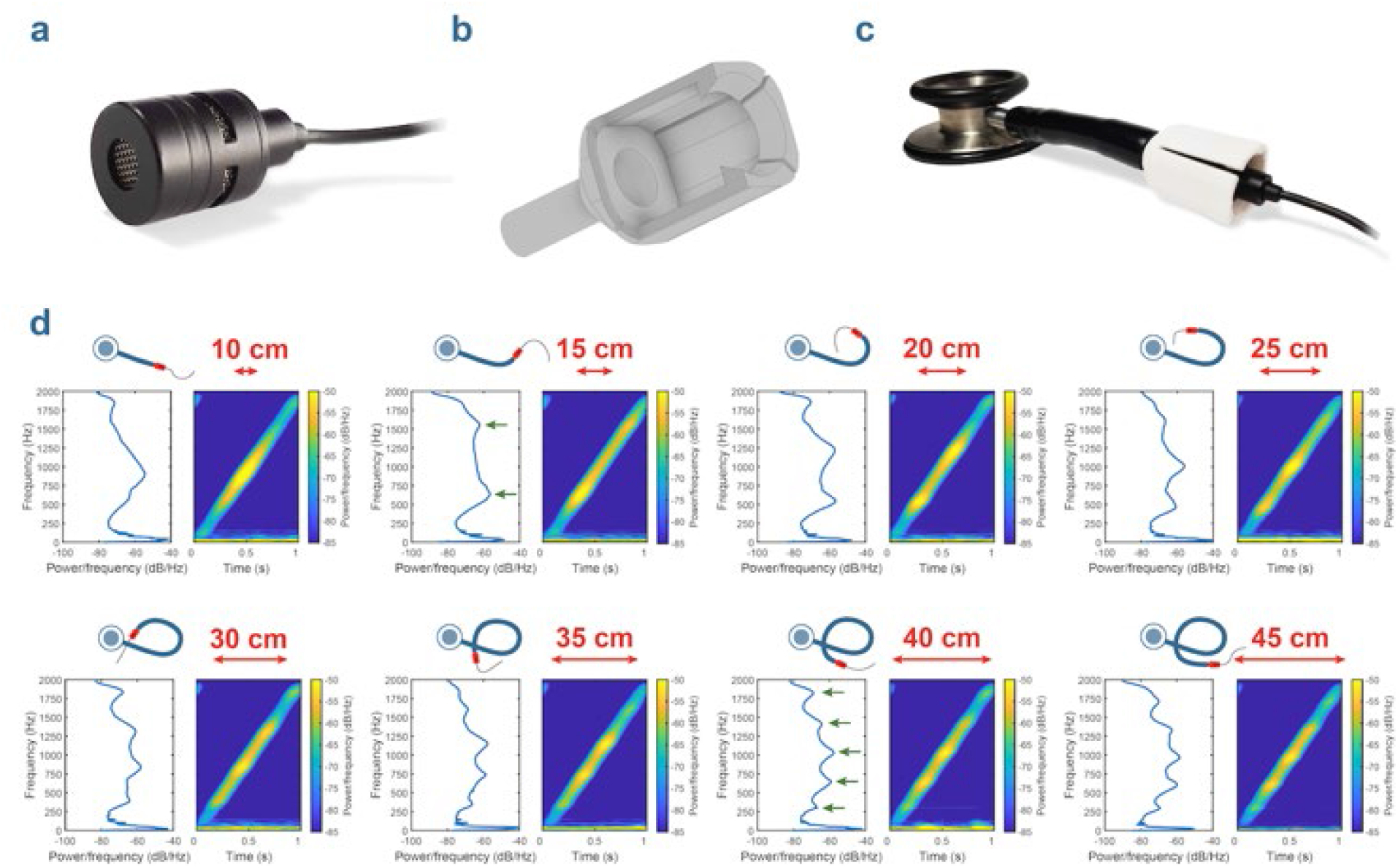
Custom made digital stethoscopes. a, SM1 Sharkoon microphone we used for the digital stethoscopes. b, 3D representation of the adapter which we designed to connect the SM1 microphone to the stethoscope tube. c, Fully assembled Littmann Classic III stethoscope with adapter and SM1 microphone. d, Frequency response of the stethoscopes for different tube length as depicted in the schematical stethoscopes above. Left Power Spectral Density (PSD) plot; right, spectrogram of the recorded chirp signal. For the PSD plot of the 15 cm and the 40 cm tube length the positions of resonance nodes are indicated with green arrows.

To ensure the quality of the recordings, it is important to securely attach the stethoscopes to the preferred locations on the patient’s body (Fig. 1c). In the user interface we suggest eight different locations, according to the CORSA guidelines [12] stethoscopes should be fixed with medical tape and eventually also gaze bandage to ensure close contact of the membrane with the body surface.

### Auscultation unit

The CORELSA system consists of bedside auscultation units to which up to three digital stethoscopes can be connected (Fig. 1b). The core of the auscultation unit is a Raspberry Pi 4 to which three sound cards are connected through USB (Fig. 1d). All components are mounted in a plastic case with the dimensions of 200 mm x 150 mm x 100 mm. We recommend using IP67 enclosures and connectors (see manual) to make sure the system is waterproof and can be disinfected properly if needed. We used a 3.5 mm audio jack to connect the stethoscopes, within the unit a 3.5 mm jack cable was soldered to the connector and then the connector was plugged into the sound card. Sound cards were connected to the Raspberry Pi with short USB connection cables. The sound cards recorded the sound files, 30 sec for manual measurements, 90 sec for automatic measurements with a sample-rate of 44.1 kHz. In post-processing, the audio files are resampled to a sample-rate of 10kHz in order to reduce the size of the data. Reducing the bandwidth of these audio files to 5kHz is sufficient because of the fact that lung sound data rarely exceeds frequencies of 2kHz ^13^.

### Auscultation Network

To remotely control the bedside auscultation units, we developed a piece of software that allows an external server to communicate with the bedside units, as well as to handle the recordings, to process the signals, generate spectrogram images and finally to send all these files to the server once completed. The embedded software for both the auscultation unit and the server was written in Python (Version 3.7, Python Software Foundation) to handle these tasks. Python is exceptionally well suited for developing this type of application, as one can rely on a wide range of libraries to implement the needed functional blocks.

The communication layer between the auscultation unit and the server is designed as a star-network, where the server can communicate with each auscultation unit. The auscultation units cannot interact with each other directly, as this would be an undesirable functionality in an ICU setting. Each auscultation unit was given a unique identifier so they could be easily traced and send specific commands. Exemplary commands are requested of the server for a new measurement, or new settings for signal gain, or automatic measurement interval. In order for the server to verify device operation, each auscultation unit was programmed to send a heartbeat message on a fixed schedule to the server. If such a heartbeat was not received by the server it was marked as offline, which could happen if there was a connection issue or power surge for example. The auscultation unit is based around a Raspberry Pi 4 as its embedded platform, with a Raspian-based operating system. Raspian is a Debian-derived Linux distribution, optimized for running on the Raspberry Pi. Using a Linux-based operating system in the bedside auscultation units makes sense for this type of application, as the application layer can greatly benefit from the operating-system functionality such as networking, parallelism and scheduling, filesystems/file-transfer and time management.

To make the recordings of the microphones we used the built-in software drivers of the operation system to accomplish the audio measurements of the connected microphones. We used a multi-threaded approach in our software application to connect to these software drivers and start the recordings, which made it possible to acquire recordings of the three microphones simultaneously. To handle the data transfer of the recordings from the auscultation unit to the server we make use of the Secure Copy Protocol (SCP). SCP is needed to perform safe and encrypted copy operations, which ensures patient privacy

### System interface

Once the auscultation units are connected to the network, a central node handled all traffic and provided an interface that can be used to control the units and acquire auscultations remotely. This interface to the system is implemented as a website using HTML5, CSS and Javascript, and lists all connected patients and has several useful diagnostic features.

A server-side python application acts as an arbiter between the ICU devices and the frontend application. This way, multiple instances of the frontend website can connect to the CORELSA system, allowing multiple physicians to collaborate on the recorded lung sound data. Devices are automatically registered and a patient’s name and/or patient ID can be linked to a unit. The user can decide whether periodical measurements are required for continuous monitoring, and/or if they want to execute a measurement on the spot. Once the measurement is available, the user can listen to the recording of all stethoscopes or view their respective spectrogram representation while listening. The spectrogram window has a zoom function to investigate certain spectrogram features or artifacts if needed. It is also possible to note the presence or absence of certain diagnostic sound features or write remarks and link them to the recording for future reference. These physician-provided labels can in a later phase be used to implement a machine-learning framework for automatic feature regression of malicious lung sounds.

## IV. Discussion

The CORELSA system was developed as a response to the COVID-19 pandemic in a very short timeframe of three weeks from idea inception to physical realization. Development began at the end of March, right before the major COVID-19 outbreak in Belgium. We developed this system as a support initiative for the health care system on a national as well as international level. The ratio behind the system is that through the reduction of patient-physician contact, and through creating the possibility for auscultation, patient follow-up in the intensive care units is facilitated. The very short development timeframe obviously significantly impacts certain design choices of the system. These choices are mostly notable in the hardware design of the CORELSA system. The system was designed with off-the-shelf components, as hardware design iteration cycles often can take a long time due to manufacturing delays. Off-the-shelf prototyping components are therefore an interesting choice. Furthermore, these off-the-shelf components increase the reproducibility of the system, as the components we used are ubiquitously available on a global level. We believe that these advantages of reproducibility and availability outweigh the disadvantages of slightly higher BOM (Bill Of Material) cost as well as feature excess. The Corelsa Hardware (BOM, enclosures, mechanical CAD files for manufacturing) is Open Hardware and is licensed under the CERN-OHLS v2, allowing redistribution and modification of the documentation and making products using it.

In addition to working with off-the-shelf components, we chose to open-source all the software needed to operate the CORELSA system. The major software components are the software for the bedside units, the software for the server backend and the frontend website application. All of these components are made available through industry-standard software containers (Docker), which allows easy reproduction of the system on similar hardware as we have used in our design. As an underlying operating system all the devices use Linux, and all the software is written in Python, which ensures cross-platform compatibility. The software is available through an open Github server, found under REF. The Corelsa Software is free software: you can redistribute it and/or modify it under the terms of the GNU General Public License as published by the Free Software Foundation, either version 3 of the License, or (at your option) any later version.

There are already other stethoscopes on the market however, they are either not scalable and expensive and none of them are open source. The suggestion to replace stethoscopes with ultrasound pocket devices during the current COVID-19 pandemic to reduce the risk of infections might also only be an option for well-equipped hospitals [9]. There are some studies descripting how to build remote stethoscopes out of low-cost components, however none of these studies gives exact tutorials to rebuild the system and none provides a framework where multiple stethoscopes and auscultation units could be controlled via a central computer [2,14-18]. The advantage of our proposed system is that it is fully documented, easy to assemble and made out of parts that should be worldwide available in large quantities.

### Clinical implications

The most important clinical implications of CORELSA have been emphasized previously. Firstly, the system diminishes the risk of contamination by reducing physical contact between COVID-19 patients and health-care workers. Physicians save time as multiple patients can be monitored from one central computer and less time is spent on putting on and taking off extensive protective gear. In addition, records are stored, allowing physicians to reanalyze data and visualize lung sounds to detect adventitious sounds. Recordings can be compared over time, which enables physicians to monitor disease progression more accurately. The latter advantages demonstrate that the application of CORELSA is not only relevant during the current COVID-19 pandemic but has the potential to be used in a wider ICU population as well.

Using the CORELSA system recordings of lung sounds are annotated by physicians, which leads to the build-up of an extensive labelled database to train machine learning algorithms. In the long-term, these algorithms may provide health-care workers with objective information on the presence and characteristics of adventitious lung sounds. This automated interpretation reduces the subjectivity of the process and could support physicians in optimizing the treatment of the patient.

### Current limitations

There are several aspects that can be improved in the current layout of our system. While we used a tube length of 45 cm for the digital stethoscopes in our prototype, measurements of different tube lengths showed that a shorter tube length of 15 cm had better transmission properties and their frequency response was comparatively flat (Fig. 2 d). For almost all tube lengths the frequency response shows a drop-off of around 15dB in between 500 Hz and 250 Hz, which is a quite important frequency range for lung sounds. Normal frequency range of pulmonary sounds varies from 100 to 1000 Hz, sometimes wheezing sounds can go up to 5000 Hz, fine crackles occur at around 650 Hz, coarse crackle at 350 Hz and rhonchi at around 150 Hz [19]. As we don’t see any attenuation of normal lung sounds in the spectrograms shown in Fig. 3 c, d, the frequency drop-off in between 500 Hz and 250 Hz is probably mainly caused by limitations of the loudspeaker we used to excite the stethoscope membrane. If they would be caused by the SM1 microphone itself however, software-based corrections of the frequency response could improve the sound quality of the system.

**Fig. 3.**
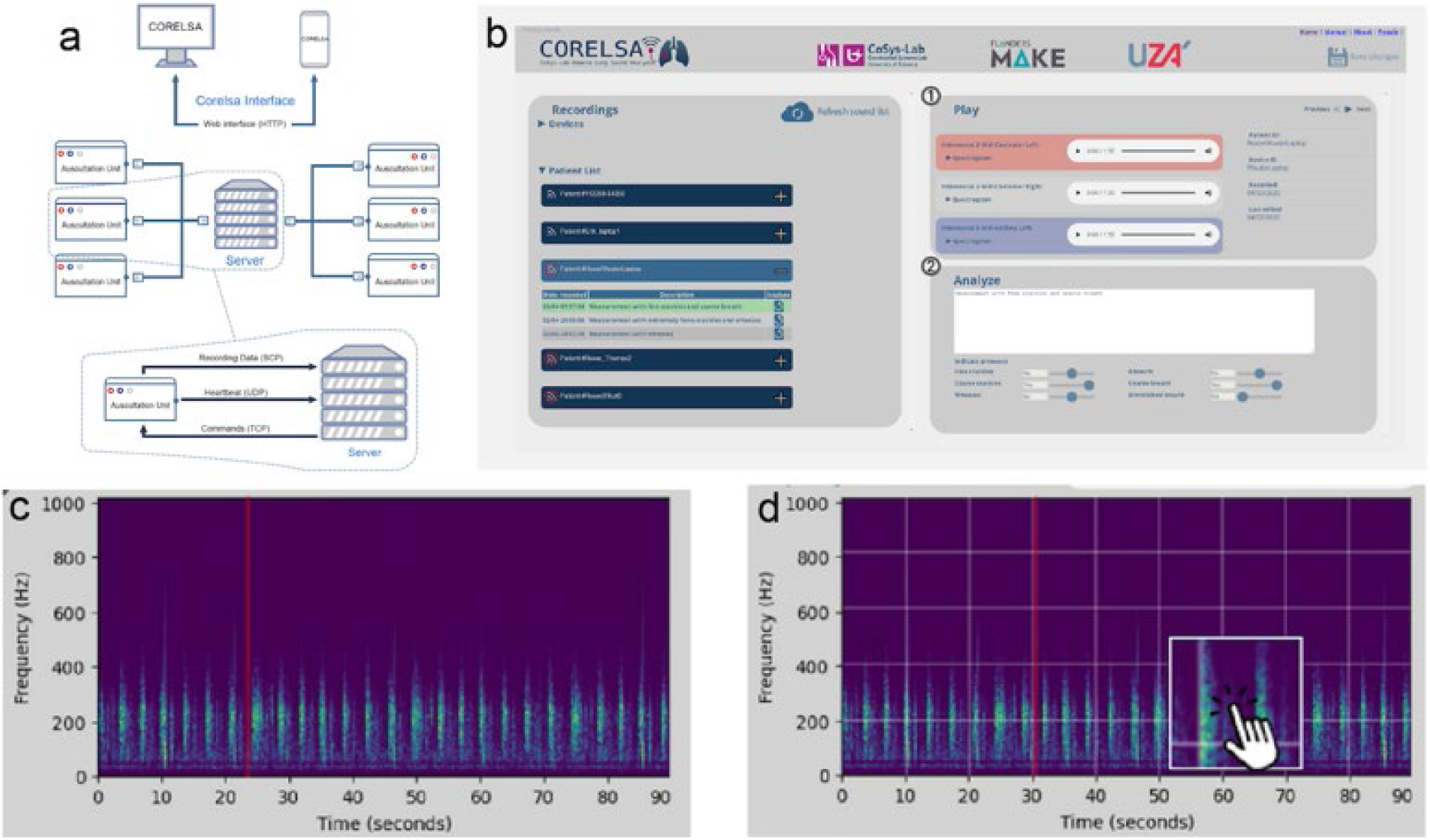
Network, interface and recordings of the CORELSA system. **a**, Overview of the network **b**, Webpage interface with which all connected auscultation units can be controlled, patient lists can be created, and auscultation recordings can be acquired. On the right, the recordings can be played and analyzed. **c**, Spectrogram of an auscultation recording of lung sounds of the first author made with the CORELSA system **b**, Spectrogram of lung sounds of the same person made directly after the recording in c with a Thinklabs digital stethoscope.

### Outlook

For our system we used Littman Classic III stethoscopes, which are comparatively expensive, nevertheless, our system could also be used with other types of stethoscopes from different manufacturers. Even 3D printed stethoscopes, like the one introduced by the Glia project [20] could be used, which would significantly reduce the costs of our system. Since the stethoscopes in our setup are constantly attached to the patients’ body, it might be uncomfortable for the patient after some time, especially because the stethoscope head is quite big. Therefore, the development of micro stethoscopes based on MEMS microphones and small diaphragms that can easily be attached to the body of the patient - just like attachable ECG Electrodes - would significantly advance our system.

In conclusion: A remote stethoscope system like CORELSA enables to perform longitudinal follow-up studies of patient lung pathologies through lung sound recording. Patient follow-up can benefit from this, as a system such as CORELSA can serve as an essential tool in the evaluation of these lung pathology developments, without significant requirements on clinician effort

## Data Availability

All data is available online at https://cosysgit.uantwerpen.be/Project/corelsa-opensource

## Notes

### Competing Interest Statement

The authors have declared no competing interest.

### Funding Statement

The work was sponsored by Flanders Make VZW, Non-Profit Strategic Research Institute, Oude Diestersebaan 133, 3920 Lommel, Belgium.

### Author Declarations

Ethics Committee of University Hospital Antwerp (UZA/UAntwerp) gave Ethical Approval for this work under number B3002020000070 The Ethics Committee of UZA/UAntwerp is a fully recognized ethics committee in accordance with the law. Its operations, composition, and responsibilities are legally established in accordance with: The Royal Decree of August 12, 1994 The European Directive 2001/20 The Law of May 7, 2004, concerning Experiments on the Human Person: Articles 11, 11/1, and 11/2, as amended by the Law of March 19, 2013 Circular No. 613 of the FAMHP (Federal Agency for Medicines and Health Products) The study was approved by the central ethics committee of UZA.

